# HCMV control in allogeneic stem cell transplant recipients - an analysis of humoral and cellular players beyond antigen-specific T cells in the letermovir era

**DOI:** 10.1101/2025.04.14.25325799

**Authors:** Chris David Lauruschkat, Hannah Görge, Kerstin Knies, Benedikt Weißbrich, Lars Dölken, Carolin Köchel, Nina Imhof, Magdalena Huber, Hartmut Hengel, Hermann Einsele, Sebastian Wurster, Sabrina Kraus

## Abstract

Allogeneic hematopoietic stem cell transplant (alloSCT) recipients often experience late-onset human cytomegalovirus (HCMV) reactivations following termination of letermovir prophylaxis. Letermovir prophylaxis extends the window for protective B- and T-cell reconstitution; however, our understanding of humoral responses and their contribution to HCMV immune control remains limited.

Combining serological and flow cytometric analyses in 42 HCMV-seropositive alloSCT recipients, we herein provide the first comprehensive longitudinal (days 90-270 post-transplant) characterization of HCMV-specific humoral responses, natural killer (NK)-cell phenotypes, and γδ T cells in the letermovir era.

HCMV controllers showed predominantly IgG-driven responses, higher pre-reactivation Vδ1^+^ γδ T-cell frequencies, and stronger expansion of “memory-like” NK cells than patients with clinically significant CMV infection (csCMVi). In contrast, csCMVi patients showed delayed HCMV-specific IgG production, IgM-skewed humoral responses, and stronger post-reactivation expansion of memory B cells and Vδ1^+^ γδ T cells. Early γδ T-cell reconstitution by day 90 was predictive of future HCMV control. HCMV-specific IgG levels correlated only weakly with γδ T cells but showed distinct associations with “memory-like” NK-cell reconstitution in HCMV controllers, suggesting synergisms between humoral and cellular immunity.

Collectively, these findings highlight a need to study anti-HCMV immune protection beyond type-1 T cells and refine risk stratification models in alloSCT patients by inclusion of novel immune markers such as γδ T-cell frequencies and phenotypes. Leveraging the extended B-cell reconstitution window created by letermovir, novel immunotherapies (e.g., therapeutic antibodies) and future vaccines might boost humoral anti-HCMV immunity and benefit from synergisms with γδ T cells and “memory-like” NK cells in improving HCMV control.

## Introduction

When conventional therapies fail, allogeneic hematopoietic stem cell transplantation (alloSCT) emerges as the most promising option for achieving a lasting cure in hematological malignancies^1^. However, alloSCT recipients are highly susceptible to opportunistic infections. Among these, reactivation of human cytomegalovirus (HCMV) represents one of the most critical complications, frequently associated with substantial morbidity and mortality^1–3^. The introduction of letermovir prophylaxis has markedly reduced early clinically significant CMV infections (csCMVi) and improved survival outcomes^4,5^. However, late-onset HCMV reactivations following letermovir cessation have become a significant clinical challenge with incidences of 25– 57% ^6–9^, underscoring the need for deeper insights into the immune mechanisms governing HCMV control.

While T cells are well-established as critical players in HCMV immunity^10–12^, the role of humoral immunity, particularly HCMV-specific B cells and antibodies, has been more scarcely studied. Historically, this was due to the slow reconstitution of B cells and the predominance of early-onset HCMV reactivations before day 100 post-transplant^13^. However, in the letermovir era, late-onset HCMV reactivations often occur when B-cell reconstitution is more advanced^14^, and antibody responses may play a crucial role in HCMV control through viral neutralization, CD16-mediated recognition of virions, antibody-dependent cellular cytotoxicity (ADCC), and cytokine secretion^15–17^.

Furthermore, other immune cell populations that are poorly explored in the letermovir era, such as “memory-like” NK cells and γδ T cells, have known anti-HCMV properties and interact with HCMV-specific antibodies via the IgG-Fc receptor CD16^15–17^, suggesting a potential interplay between humoral and cellular immunity that warrants further investigation.

To address these knowledge gaps, we conducted a longitudinal analysis of peripheral blood mononuclear cell (PBMC) and serum samples from 42 HCMV-seropositive alloSCT patients at four post-transplantation time points: day 90 (prior to letermovir cessation), day 150 (around the median time of first HCMV reactivation), and days 210 and 270 (following HCMV reactivation). We compared patients who controlled HCMV reactivation with those who developed csCMVi to investigate the roles of humoral anti-HCMV responses, “memory-like” NK cells, and γδ T cells in HCMV defense (**Figure 1A**). Thereby, we provide novel insights into the immune mechanisms underlying HCMV control in the letermovir era beyond antigen-specific T cells, paving the way for improved strategies in risk stratification, vaccination, and immunotherapy development.

**Figure 1.**
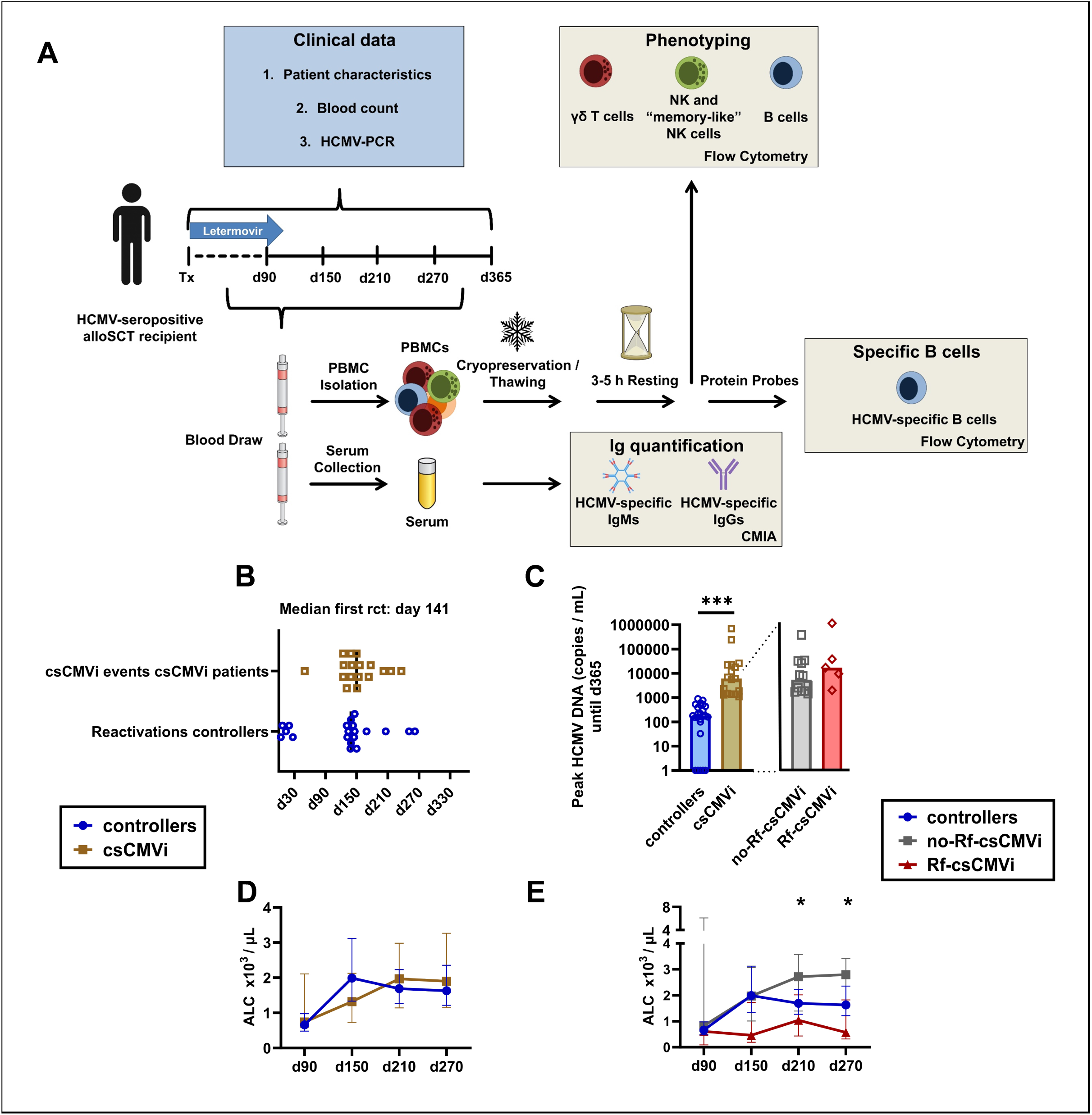
HCMV reactivation timing, HCMV viral load, and absolute lymphocyte counts. (**A**) Study flowchart. (**B**) Median day of first HCMV reactivation in controllers (N = 25) or onset of csCMVi in affected patients (N = 17). No HCMV viral load was detected in five controllers throughout the observation period. (**C**) Peak HCMV DNA copies/mL until day 365 (measured by PCR). csCMVi patients were further subdivided into those without (grey) or with Rf-csCMVi (red). Mann-Whitney U test. (**D-E**) ALC kinetics for controllers and csCMVi patients (Mann-Whitney U test) (**D**), as well as controllers, no-Rf-csCMVi, and Rf-csCMVi patients. Kruskal-Wallis (**E**). * p < 0.05, *** p < 0.001. <u>Abbreviations</u>: ALC = absolute lymphocyte counts, csCMVi = clinically significant HCMV infection, HCMV = human cytomegalovirus, PCR = polymerase chain reaction, Rf-csCMVi = refractory clinically significant HCMV infection.

## Materials and Methods

### Institutional review board approval

This study was approved by the Ethics Committees of the University of Würzburg, Germany (protocol code 17/19-sc). Written informed consent was obtained from all patients.

### Study population

Forty-two HCMV-seropositive alloSCT recipients (Recipient [R]+Donor[D]+, R+D-) were enrolled at the University Hospital of Würzburg (Germany) between December 2019 and October 2023. All patients received letermovir prophylaxis at a dose of 480 mg once daily from day 1 to day 100 post-transplant, with a reduced dose of 240 mg once daily for those receiving cyclosporine A. HCMV DNAemia was monitored weekly until day 100 post-transplant and biweekly thereafter using real-time PCR. If HCMV DNA levels exceeded 1,000 copies/mL, letermovir prophylaxis was discontinued, and preemptive systemic antiviral therapy with (val)ganciclovir was initiated. Additionally, cryopreserved PBMCs and serum samples from 15 patients on preemptive therapy without letermovir prophylaxis, collected between September 2015 and June 2019, were used for supplementary analyses.

### Definitions

Based on a follow-up period until day 365 post-transplant, subjects were categorized as HCMV controllers or csCMVi patients. The csCMVi group included patients who required antiviral therapy due to HCMV DNA blood levels exceeding 1,000 copies/mL, whereas those who maintained HCMV DNA levels <1,000 and did not require antiviral treatment were classified as HCMV controllers. For some analyses, the csCMVi group was further subdivided into patients with probable refractory HCMV infection (Rf-csCMVi), as defined by Chemaly et al.^18^, and those without refractory infection.

For some analyses, cell population counts were compared before and after csCMVi. Pre-csCMVi measurements were taken at the last available time point prior to the initiation of anti-HCMV therapy. Post-csCMVi assessments were performed using samples collected at the earliest available time point when HCMV DNA levels had returned to <300 copies/mL. In controllers, pre-reactivation measurements were taken before HCMV DNA levels ranged between 1 and 1,000, while post-reactivation measurements were obtained once levels returned to zero.

### Immunoassays

Ethylenediaminetetraacetic acid-anticoagulated blood was collected on days 84-106 (day 90), 141-167 (day 150), 200-233 (day 210), and 262-309 (day 270) post-transplant to isolate and cryopreserve PBMCs for flow cytometric analysis of γδ T-, B-, and NK-cell phenotypes and HCMV-specific B cells. HCMV-specific B cells were detected using biotinylated protein probes bound to streptavidin fluorophores. Frequencies of immune cell subpopulations were calculated using a combination of flow cytometry data and absolute lymphocyte counts. Detailed protocols and gating strategies are provided in the **Supplementary Material**. Serum samples were analyzed for HCMV-specific IgM and IgG using the Architect CMV IgG and IgM kits (Abbott Ireland), following the manufacturer’s instructions.

### Statistical analysis

Significance testing was performed using the Mann-Whitney U test, paired Wilcoxon test, Kruskal-Wallis test, or Fisher’s exact test, as appropriate. Data were compiled, visualized, and analyzed using Microsoft Excel (Microsoft, Redmond, WA, USA), Prism v.10.2 (GraphPad Software, Boston, MA, USA), and R v.4.2.1 (R Core Team).

## Results

### Patient characteristics

Forty-two HCMV-seropositive alloSCT patients who received letermovir prophylaxis during the first 100 days post-transplantation were included in this study. Among them, 25 patients (60%) successfully controlled HCMV infection until day 365 post-transplant, whereas 17 patients (40%) experienced csCMVi. Among those with csCMVi, 5 patients had Rf-csCMVi. The median time of first HCMV reactivation was day 141, with no significant difference (p=0.266) in timing between the non-clinically significant HCMV reactivations in controllers (median, day 139) and csCMVi events in the csCMVi group (median, day 150) (**Figure 1B**). Expectedly, the median peak HCMV DNA copies until day 365 were significantly higher in csCMVi patients than in controllers (6,100 copies/mL vs. 180 copies/mL, p=<0.001), with no significant difference observed between the Rf-csCMVi and non-Rf-csCMVi subgroups (**Figure 1C**).

A detailed breakdown of patient characteristics is provided in **Table 1**. Demographics and underlying patient characteristics did not differ significantly between controllers and csCMVi patients, except for a higher incidence of 2^nd^ or higher-grade acute graft-versus-host disease (aGvHD) in csCMVi patients (p=0.018, **Table 1**). To preclude a significant impact of pharmacologic immunosuppression on our immunologic analyses, we specifically analyzed the number and dosage of immunosuppressants administered during and after letermovir prophylaxis. A slightly higher proportion of csCMVi patients than controllers (18% vs. 8%) received systemic high-dose glucocorticosteroids (GCS), i.e., prednisolone at a daily dose of 2 mg/kg for GvHD treatment after letermovir cessation. However, second- or third-line therapies for GvHD with non-GCS immunosuppressive agents were similar between the two cohorts (**Table 2**).

**Table 1.** Characteristics of alloSCT patients who successfully controlled HCMV (controllers) and those who developed clinically significant HCMV infection (csCMVi). Mann–Whitney U test or Fisher’s exact test was applied as appropriate. <u>Abbreviations</u>: aGvHD = acute graft versus host disease, cGvHD = chronic graft versus host disease, HCT-CI = hematopoietic cell transplantation comorbidity index, HLA = human leukocyte antigen, PBSC = peripheral blood stem cells, R/D = recipient / donor.

| **Variables** | **Total**<br>**(n=42)** | **HCMV controllers**<br>**(n=25)** | **csCMVi**<br>**(n=17)** | **p\*** |
| --- | --- | --- | --- | --- |
| **Age, median (range)** | 62 | 62 (22-73) | 61 (33-71) | 0.995 |
| **Sex, n (%)** |  |  |  |  |
| Male | 21 (50) | 14 (56) | 7 (41) | 0.530 |
| Female | 21 (50) | 11 (44) | 10 (59) |  |
| **Underlying disease, n (%)** |  |  |  |  |
| Chronic leukemia | 1 (2) | 1 (4) | 0 (0) | - |
| Multiple myeloma | 2 (5) | 1 (4) | 1 (6) |  |
| Acute leukemia | 25 (60) | 14 (56) | 11 (65) |  |
| Lymphoma | 0 (0) | 0 (0) | 0 (0) |  |
| Others | 14 (33) | 9 (36) | 5 (29) |  |
| **HLA matching, n (%)** |  |  |  |  |
| Matched related | 5 (12) | 1 (4) | 4 (24) | - |
| Matched unrelated | 24 (57) | 19 (76) | 5 (29) |  |
| Haploidentical | 0 (0) | 0 (0) | 0 (0) |  |
| Mismatch | 13 (31) | 5 (20) | 8 (47) |  |
| **Stem cell source, n (%)** |  |  |  |  |
| Bone marrow | 1 (2) | 1 (4) | 0 (0) | >0.999 |
| PBSC | 41 (98) | 24 (96) | 17 (100) |  |
| **Conditioning Regimen, n (%)** |  |  |  |  |
| Reduced intensity (RIC) | 39 (93) | 23 (92) | 16 (94) | >0.999 |
| Myeloablative (MAC) | 3 (7) | 2 (8) | 1 (6) |  |
| **Antithymocyte globulin, n (%)** |  |  |  |  |
| No | 5 (12) | 1 (4) | 4 (24) | 0.140 |
| Yes | 37 (88) | 24 (96) | 13 (76) |  |
| **HCT-CI, n (%)** |  |  |  |  |
| 0-2 | 30 (71) | 19 (76) | 11 (65) | - |
| 3-4 | 9 (21) | 4 (16) | 5 (29) |  |
| ≥5 | 3 (7) | 2 (8) | 1 (6) |  |
| **aGvHD, n (%)** |  |  |  |  |
| 0-1 | 29 (69) | 21 (84) | 8 (47) | 0.018* |
| 2-4 | 13 (31) | 4 (16) | 9 (53) |  |
| **cGvHD, n (%)** |  |  |  |  |
| No | 27 (64) | 16 (64) | 11 (65) | >0.999 |
| Yes | 15 (36) | 9 (36) | 6 (35) |  |
| **HCMV serostatus (R/D), n (%)** |  |  |  |  |
| +/+ | 26 (62) | 17 (68) | 9 (53) | 0.353 |
| +/- | 16 (38) | 8 (32) | 8 (47) |  |
| **One-year mortality, n (%)** |  |  |  |  |
| Alive | 35 (83) | 21 (84) | 14 (82) | >0.999 |
| Dead | 7 (17) | 4 (16) | 3 (18) |  |

**Table 2.** Immunosuppressant administration during and after letermovir prophylaxis in controllers and csCMVi patients. <u>Abbreviations</u>: csCMVi = clinically significant HCMV infection (csCMVi), LET = Letermovir

|  | **Total** |  | **HCMV controllers** |  | **csCMVi** |  |
| --- | --- | --- | --- | --- | --- | --- |
|  | **on Let** | **after**<br>**LET** | **on LET** | **after**<br>**LET** | **on LET** | **after**<br>**LET** |
| **Steroid use\*** |  |  |  |  |  |  |
| None | 36 (86) | 37 (88) | 24 (96) | 23 (92) | 12 (71) | 14 (82) |
| >0 mg to ≤1 mg/kg/d | 2 (5) | 0 (0) | 1 (4) | 0 (0) | 1 (6) | 0 (0) |
| 2 mg/kg/d | 4 (10) | 5 (12) | 0 (0) | 2 (8) | 4 (24) | 3 (18) |
| **Immunosuppressants<sup>#</sup>** |  |  |  |  |  |  |
| 0 | 0 (0) | 39 (93) | 0 (0) | 23 (92) | 0 (0) | 16 (94) |
| 1 | 0 (0) | 0 (0) | 0 (0) | 0 (0) | 0 (0) | 0 (0) |
| 2 | 40 (95) | 2 (5) | 25 (100) | 2 (8) | 15 (88) | 0 (0) |
| > 2 | 2 (5) | 1 (2) | 0 (0) | 0 (0) | 2 (12) | 1 (6) |

\* Divided into 0, >0 to ≤1, or 2 mg/kg/d based on maximum steroid dose during the study period.
<sup>#</sup> Not including steroids (e.g. ruxolitinib, tacrolimus, sirolimus etc.)

While absolute lymphocyte counts (ALC) values were similar in controllers and csCMVi patients, significant differences emerged in 3-group comparisons between controllers, Rf-csCMVi, and non-Rf-csCMVi from day 210 onward. Specifically, patients with Rf-csCMVi patients had significantly lower median day-210 (1.0 vs. 1.7 vs. 2.7 × 10³/µL, p=0.030) and day 270 (0.6 vs. 1.6 vs. 2.8 × 10³/µL, p=0.0177) ALCs than both controllers and non-Rf-csCMVi patients, respectively (**Fig. 1D-E**).

### AlloSCT recipients in the letermovir era mount robust humoral responses by the time of HCMV reactivation and develop memory B cells after reactivation events

Before the introduction of letermovir, there was no routine prophylaxis against HCMV. As a result, humoral immunity played a limited role in protection, primarily due to the predominance of early HCMV reactivations and slow reconstitution of B cells^11^. Analyses of cryopreserved patient samples from the pre-letermovir era revealed a lack of B-cell reconstitution by the time of HCMV reactivation (median, day 26 post-transplant) (**Figure 2A**). Consequently, longitudinal analysis of serum samples from the same patients showed no increase in HCMV-specific IgG or IgM from day 30 to day 120, indicating a lack of a functional humoral response to HCMV reactivation events (**Figure 2B**).

**Figure 2.**
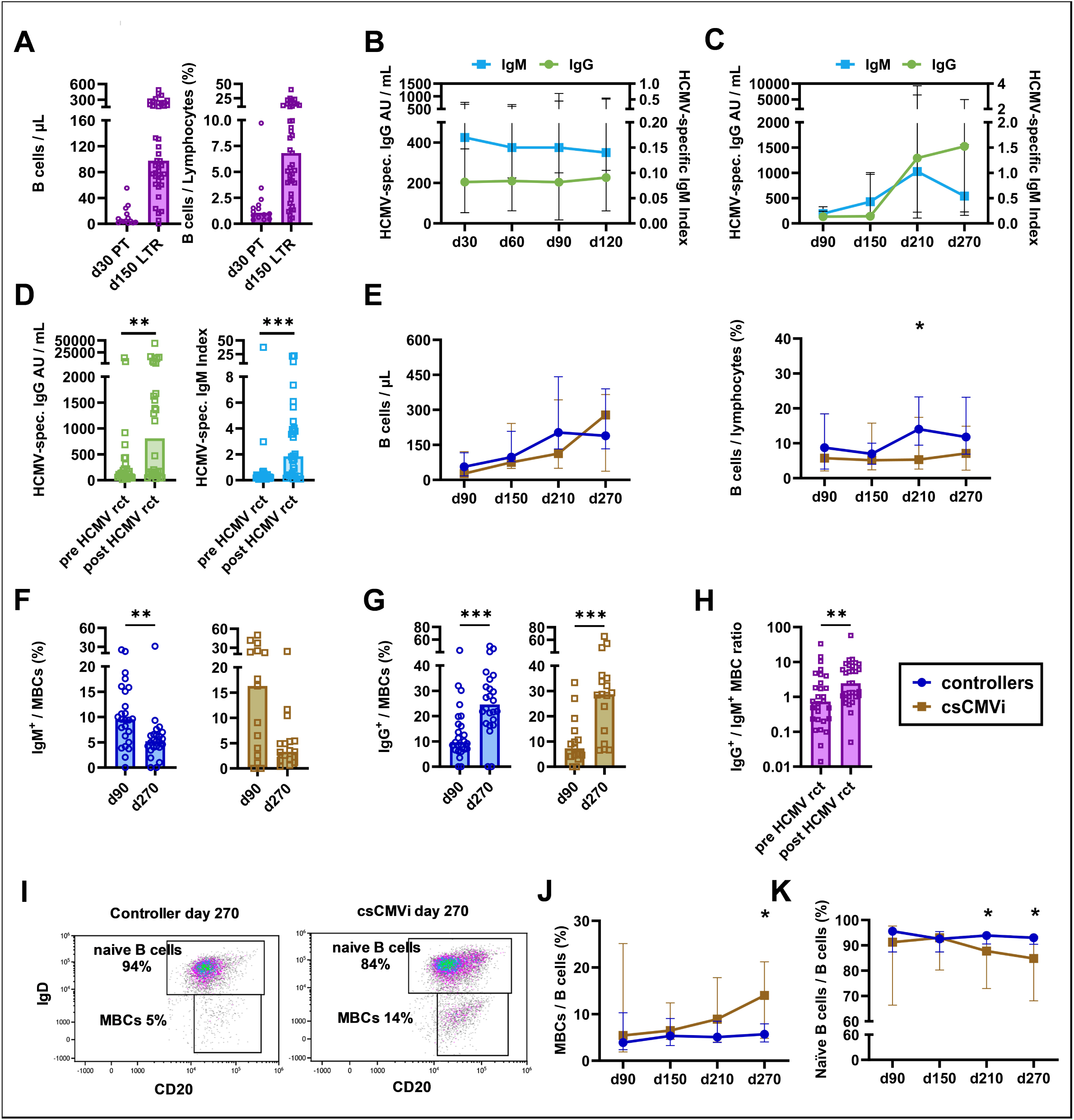
HCMV reactivation is associated with a robust humoral response and memory B-cell formation. Samples from alloSCT patients receiving letermovir prophylaxis (N = 42) or historic samples from the pre-letermovir era (N = 15, A and B only) were analyzed using flow cytometry for cellular markers and CMIA for serological markers. (**A**) B-cell counts and frequencies around the median time of first HCMV reaction in the pre-letermovir era (day 30) and in patients receiving letermovir prophylaxis (day 150). Mann-Whitney U test. (**B–C**) Kinetics of HCMV-specific IgM and IgG levels in the pre-letermovir era (**B)** and in patients receiving letermovir prophylaxis with preemptive therapy around the time of HCMV reactivation (**C-D**). (**E**) B-cell counts and frequencies in controllers and csCMVi patients in the letermovir era. Mann-Whitney U test. (**F–G**) Evolution of IgM⁺ (**F**) and IgG⁺ (**G**) MBC frequencies between day 90 and day 270 in controllers and csCMVi patients. Paired Wilcoxon test. (**H**) Ratio of IgG⁺/IgM⁺ MBC frequencies before and after HCMV reactivation. Paired Wilcoxon test. (**I**) Representative flow cytometry plot showing differentiation of naïve B cells and MBCs. (**J–K**) Kinetics of MBC (**I**) and naïve B-cell (**J**) frequencies in controllers and csCMVi patients. Mann-Whitney U test. p < 0.05, ** p < 0.01, *** p < 0.001. <u>Abbreviations:</u> alloSCT = allogeneic stem cell transplantation, CMIA = chemiluminescent microparticle immunoassay, csCMVi = clinically significant CMV infection, d = day, HCMV = human cytomegalovirus, Ig = immunoglobulin, LVR = letermovir, MBCs = memory B cells, rct = reactivation, PT = preemptive therapy.

In contrast, in our current cohort from the letermovir era, HCMV reactivation occurred significantly later (median, 141). At this later time point, we expectedly observed more readily reconstituted B-cell numbers (median, 97.5) and frequencies (median, 6.8% of total lymphocytes) (**Figure 2A**). Furthermore, HCMV-specific IgG and IgM increased between days 150 and 210 (**Figure 2C**), i.e., around the time of HCMV reactivation in most patients (**Figure 1B**). Comparisons of pre- and post-reactivation measurements further confirmed significant expansion of HCMV-specific IgG (131 vs. 809 AU/mL, p=0.009) and IgM (0.24 vs. 1.84 IgM Index, p=<0.001) (**Figure 2D**).

Interestingly, total B-cell numbers and frequencies of csCMVi patients and controllers were largely comparable, except for higher B-cell frequencies in controllers at day 210 (**Figure 2E**). In both groups, IgM^+^ B cells decreased from day 90 to day 270, whereas IgG^+^ B cells increased, consistent with expected patterns of immune reconstitution and memory formation including antibody class switching (**Figure 2F-G**). Furthermore, the ratio of IgG^+^ to IgM^+^ memory B cells (MBCs) increased from pre- to post-HCMV events (p=0.006, **Figure 2H**), a trend that was observed in both controllers (p=0.040) and csCMVi patients (p=0.085).

Although global B-cell frequencies were largely comparable, the time-dependent evolution of B-cell phenotypes differed significantly between HCMV controllers and csCMVi patients. Specifically, those with csCMVi displayed significantly higher day-270 frequencies of MBCs (14.0 vs. 5.7% among B cells, p=0.032) and lower frequencies of naïve B cells (84.8 vs. 93.0% among B cells, p=0.018) than controllers (**Figure 2I-K**).

In summary, alloSCT patients in the letermovir era mount robust HCMV-specific humoral responses by the time of HCMV reactivation, suggesting a potential novel role of the humoral response in anti-HCMV immunity. Specifically, the time-dependent shift in B-cell phenotypes gives rise to the hypothesis that HCMV reactivations may shape the B-cell repertoire in these patients.

### HCMV reactivation triggers the expansion of HCMV-specific B cells

To test this hypothesis, we longitudinally analyzed the magnitude and quality of HCMV-specific humoral response in our letermovir cohort in more detail. Therefore, HCMV-specific MBCs were detected using fluorescent protein probes targeting three key HCMV-derived antigens eliciting neutralizing antibody responses, the gH/gL/gO trimer, the gH/gL/UL128/UL130/UL131A pentamer, and glycoprotein B (gB) (**Figure 3A-B**). Quantifying these antigens individually and in combination showed high specificity, with no HCMV-specific MBCs detected in HCMV-seronegative alloSCT recipients (**Figure S1A**).

**Figure 3.**
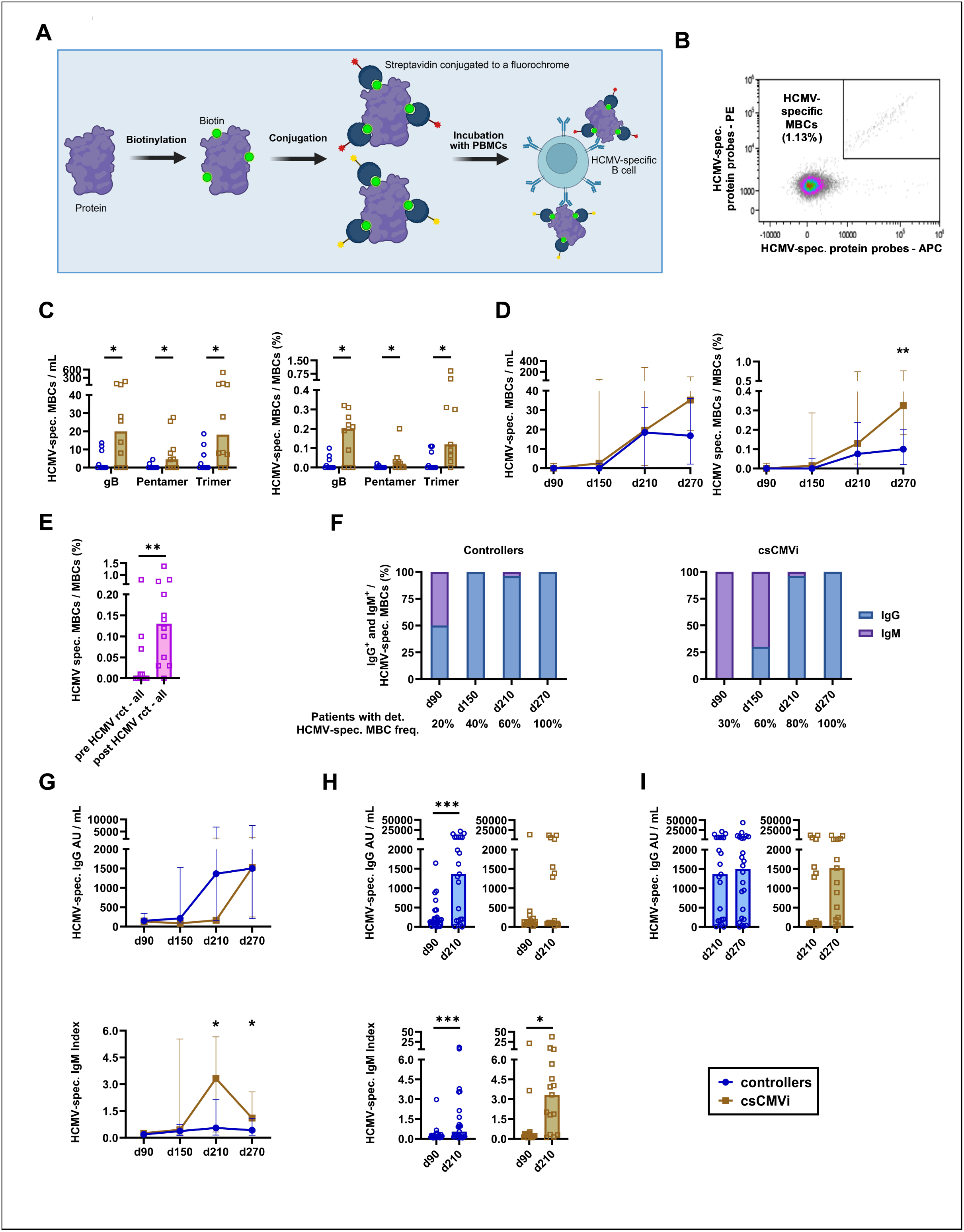
csCMVi patients show an IgM-skewed HCMV-specific response with delayed IgG kinetics compared to controllers. Reconstitution of the HCMV-specific humoral immune response was analyzed by flow cytometry and CMIA. (**A**) Flowchart illustrating the generation of HCMV-specific protein probes to detect HCMV-specific B cells. (**B**) Representative flow cytometry plot showing the detection of HCMV-specific MBCs. (**C**) Absolute numbers and frequencies of HCMV-specific MBCs targeting glycoprotein B (gB), the pentamer complex (gH/gL/UL128/UL130/UL131A), and the trimer complex (gH/gL/gO) on day 270. N = 20. Mann-Whitney U test. (**D**) Kinetics of absolute numbers and frequencies of combined gB-, pentamer-, and trimer-specific MBCs. N = 20. Mann-Whitney U test. (**E**) HCMV-specific MBC frequencies before and after HCMV reactivation in patients with detectable HCMV viral load. N = 12. Paired Wilcoxon test. (**F**) Normalized median frequencies of IgG^+^ and IgM^+^ HCMV-specific MBCs over time. N = 20. (**G**) Kinetics of HCMV-specific IgG levels and IgM indices. N = 40. Mann-Whitney U test. (**H-I**) Evolution of serologic responses between days (**H**) 90 and 210 and (**I**) day 210 and 270, respectively. N = 40. Paired Wilcoxon test. *p < 0.05, **p < 0.01, ***p < 0.001. <u>Abbreviations:</u> alloSCT = allogeneic stem cell transplantation; CMIA = chemiluminescent microparticle immunoassay; csCMVi = clinically significant CMV infection; d = day; HCMV = human cytomegalovirus; Ig = immunoglobulin; MBCs = memory B cells; rct = reactivation.

Compared to controllers, higher counts and frequencies of gB-, trimer-, and pentamer-specific MBCs were found in csCMVi patients on day 270 (**Figure 3C**). Given the low frequencies of HCMV-specific MBCs early after transplantation and limited cell availability, we combined the probes for all three antigens for subsequent longitudinal quantification. This approach revealed strongly increasing HCMV-specific MBCs in both HCMV controllers and csCMVi patients between days 90 and 210. However, by day 270, csCMVi patients had developed higher HCMV-specific MBC counts (35.1 vs. 16.8, p=0.064) and frequencies (0.33% vs. 0.10%, p=0.009) than controllers (**Figure 3D)**. To determine whether the increase in HCMV-specific MBCs resulted from HCMV reactivation, we further compared their frequencies before and after reactivation in patients with detectable HCMV viral load, regardless of csCMVi status. Indeed, HCMV-specific MBC frequencies significantly increased from a median of 0.00% pre-reactivation to 0.13% post-reactivation (p=0.001) (**Figure 3E**). Overall, these results suggest that HCMV reactivation drives HCMV-specific MBCs development in seropositive alloSCT patients.

### csCMVi patients display an IgM-skewed HCMV-specific response during reactivation and a delayed IgG response compared to controllers

Given the disparate affinity and antiviral effector responses of IgG and IgM^19^, we next examined differences in HCMV-specific IgG and IgM serum concentration in controllers and csCMVi patients. On day 150, i.e., around the median time of HCMV reactivation (**Figure 1B**), 100% of the detected HCMV-specific MBCs in controllers showed an IgG phenotype. In contrast, csCMVi patients predominantly exhibited an IgM phenotype (70% IgM^+^, 30% IgG^+^) (**Figure 3F**).

These differences were also reflected in longitudinal serum levels of HCMV-specific IgG and IgM. HCMV controllers showed a non-significant trend toward higher HCMV-specific IgG levels on day 150 (median 212 vs. 82 AU/mL, MMR=2.6) and day 210 (median 1363 vs. 163 AU/mL, MMR=8.4). Conversely, csCMVi patients had significantly higher HCMV-specific IgM indices on day 210 (median 3.3 vs. 0.6, MMR=5.5, p=0.029) and day 270 (1.1 vs. 0.4, MMR=2.8, p=0.046) (**Figure 3G**).

Moreover, controllers showed a robust increase in HCMV-specific IgG levels between days 90 and 210 (147 vs. 1363 AU/mL, p=<0.001), but only a small increase in HCMV-specific IgM indices (0.2 vs. 0.6, p=<0.001). Inversely, csCMVi patients mounted a strongly increasing HCMV-specific IgM response during that timeframe (0.3 vs. 3.3, p=0.015) but showed no significant increase in HCMV-specific IgG levels (130 vs. 164 AU/mL, p=0.421) (**Figure 3H**). Instead, csCMVi patients had a delayed rise in HCMV-specific IgG levels compared to controllers that occurred between day 210 and day 270 (163 vs. 1523 AU/mL, median fold change=9.3), resulting in comparable IgG levels between the two groups by day 270 (median 1523 vs. 1500 AU/mL) (**Figure 3I**).

Overall, time-dependent HCMV-specific B cell responses differed significantly between controllers and csCMVi patients. Controllers displayed modest, early IgG-tilted development, whereas csCMVi patients exhibited a comparatively more IgM-skewed humoral response upon viral reactivation. This suggests a delayed IgM-to-IgG class switch in csCMVi patients.

### csCMVi events drive the expansion of Vδ1^+^ γδ T cells and shift the Vδ1^+^/Vδ2^+^ ratio

In addition to directly neutralizing virions, HCMV-specific IgG can enhance other determinants of immune defense^15–17^. Specifically, immune cells with anti-HCMV properties, including γδ T cells and memory-like NK cells, can interact in part with HCMV-specific IgGs via the IgG receptor CD16^15–17^. To investigate the potential correlations between humoral and cellular immunity, we characterized the reconstitution of these immune cell populations.

Firstly, we evaluated the reconstitution of γδ T cells, a cell population that is protective against HCMV reactivation in alloSCT patients (**Figure 4A**)^20–22^. Both controllers and csCMVi patients had similar γδ T-cell kinetics and significantly expanded their total γδ T-cell numbers from day 90 to day 210 (**Figure 4B-C**). However, upon phenotypic analysis (**Figure 4D**), distinct sub-populations of these cells differed significantly between controllers and csCMVi patients over time. Specifically, controllers had higher Vδ1^+^ cell numbers (3.7 vs. 1.8 cells/µL, MMR=2.1) and frequencies (40.7 vs. 19.6% Vδ1+ cells among γδ T cells, p=0.050) than csCMVi patients before letermovir cessation (day 90) (**Figure 4E**). In fact, high day-90 Vδ1⁺ cell frequencies were predictive of subsequent HCMV control, as confirmed by receiver-operating characteristics (ROC) analysis (area under the curve 0.688, p=0.049, 95% confidence interval: 0.4967-0.8793, **Figure 4F**). Inversely, csCMVi patients showed higher Vδ1^+^ frequencies among γδ T cells than controllers from day 210 onward (day 210: 72.0 vs. 56.0%, p=0.032; day 270: 73.6 vs. 59.9%, p=0.021) (**Figure 4E**). Vδ2⁻ cells followed the trend of Vδ1⁺ cells (**Figure S1B-E**).

**Figure 4.**
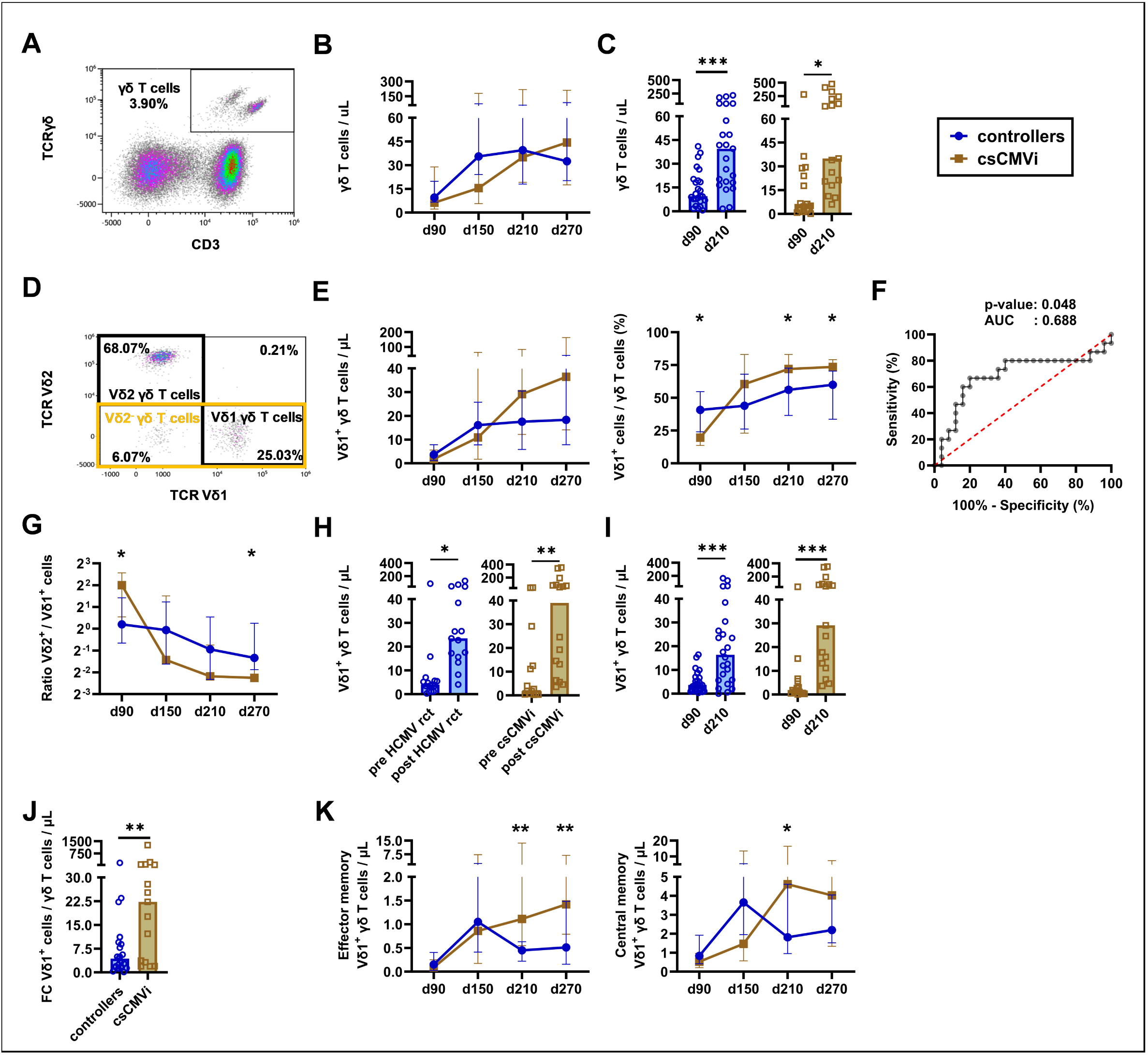
csCMVi events promote Vδ1^+^ γδ T cell expansion and alter the Vδ1^+^/Vδ2^+^ ratio. Reconstitution of γδ T cells in 42 alloSCT recipients was analyzed by flow cytometry. (**A**) Representative flow cytometry plot for quantification of γδ T cells. (**B**) Kinetics of γδ T-cell numbers. Mann-Whitney U test. (**C**) Comparison of γδ T-cell numbers on days 90 and 210. Paired Wilcoxon test. (**D**) Representative flow cytometry plot illustrating γδ T-cell subpopulations. (**E**) Kinetics of absolute numbers and frequencies of Vδ1^+^ γδ T cells. Mann-Whitney U test. (**F**) ROC analysis for prediction of csCMVi events based on Vδ1^+^ γδ T-cell frequencies on day 90. (**G**) Longitudinal assessment of the Vδ2^+^/Vδ1^+^ ratio. Mann-Whitney U test. (**H**) Quantification of Vδ1^+^ γδ T cells before and after HCMV reactivation/csCMVi events. Paired Wilcoxon test. (**I**) Comparison of Vδ1^+^ γδ T-cell counts at days 90 and 210. Paired Wilcoxon test. (**J**) Intra-individual fold changes (FC) of Vδ1^+^ γδ T-cell counts from day 90 until day 210. Mann-Whitney U test. (**K**) Kinetis of effector and central memory Vδ1^+^ γδ T-cell counts. Mann-Whitney U test. *p < 0.05, **p < 0.01, ***p < 0.001. <u>Abbreviations:</u> alloSCT = allogeneic stem cell transplantation; AUC = area under the curve; csCMVi = clinically significant CMV infection; d = day; FC = fold changes; rct = reactivation; ROC = receiver operating characteristic curve.

Vδ2^+^ frequencies showed minimal differences between HCMV controllers and csCMVi patients. Likewise, comparable numbers and frequencies of the Vδ2^+^Vδ9^+^ and Vδ2^+^Vδ9^−^ subpopulations were found in both groups (**Figure S1F-Q**).

In csCMVi patients, these trends collectively led to a marked shift in Vδ1⁺/Vδ2⁺ ratios over time, which were initially higher (day 90: Vδ2^+^/Vδ1^+^ 4.0 vs. 1.2, p=0.038) and later lower (day 270: Vδ2^+^/Vδ1^+^ 0.2 vs. 0.4, p=0.015) than in controllers (**Figure 4G**). While both groups expanded Vδ1^+^ cell numbers post-reactivation/csCMVi events, csCMVi patients had a significantly greater magnitude of expansion between day 90 and 210 than controllers (median fold changes, 22.3 vs. 4.3, p=0.007, **Figure 4H-J**).

Furthermore, differences in Vδ1⁺ memory populations were observed between the two cohorts from day 210 onward. Specifically, csCMVi patients had significantly higher numbers of effector memory (day 270: 1.4 vs. 0.5 CDRA⁻CD27⁻ Vδ1⁺ cells/μL, p=0.003) and central memory Vδ1⁺ cells (day 210: 4.6 vs. 1.8 CDRA⁻CD27⁺ Vδ1⁺ cells/μL, p=0.023) than controllers (**Figure 4K**). In contrast, naïve and terminally differentiated effector cells remained comparable (**Figure S1R-S**).

In summary, low Vδ1^+^ frequencies before letermovir cessation were predictive for future csCMVi events. csCMVi events were, in turn, associated with significant expansion of Vδ1^+^ γδ T cells, resulting in an increased Vδ1^+^/Vδ2^+^ ratio compared to controllers, partially due to expansion of effector and central memory phenotypes.

### Lack of “memory-like” NK-cell expansion is associated with refractory csCMVi

Like γδ T cells, NK cells, and especially “memory-like” NK cells, play a crucial role in controlling HCMV after alloSCT and possess proficient IgG-dependent ADCC capabilities^16,23^. Therefore, we analyzed NK-cell reconstitution in our cohort.

Consistent with prior evidence^10^, there were no differences in total NK-cell numbers or the distribution of CD56^dim^ and CD56^bright^ NK-cell subpopulations between HCMV controllers and csCMVI patients (**Figure 5A-B**). Likewise, there was no major change in the expansion of these cell populations before and after csCMVi except for a minor shift toward CD56^dim^ post-reactivation (**Figure 5C**).

**Figure 5.**
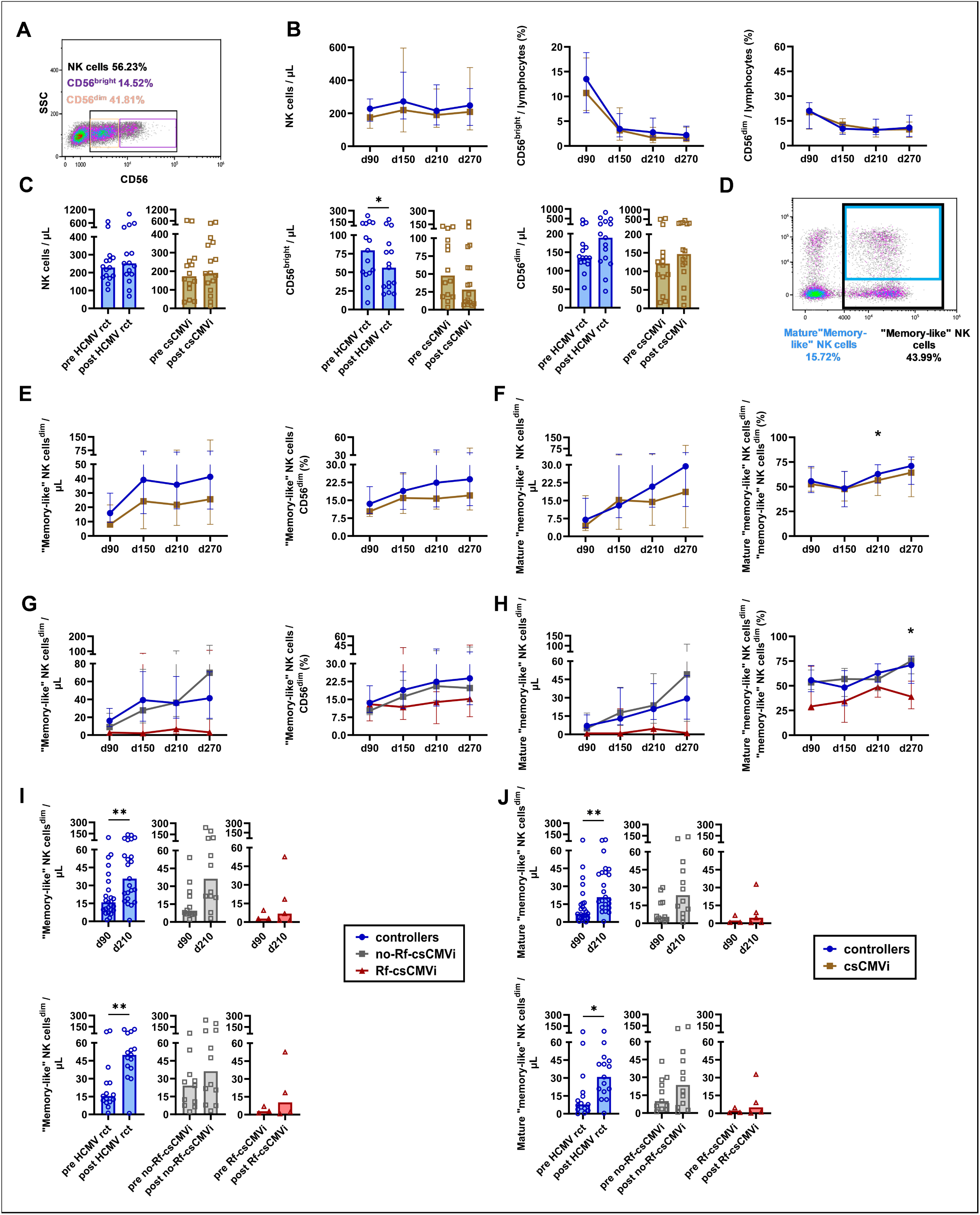
Failure to expand “memory-like” NK cells is linked to refractory csCMVi. Reconstitution of NK cells in alloSCT recipients (N = 42) was analyzed by flow cytometry. (**A**) Representative flow cytometry plot of NK-cell subpopulations. (**B**) Kinetics of total NK cells and the frequencies of CD56^dim^ and CD56^bright^ NK-cell subpopulations. Mann-Whitney U test. (**C**) Absolute numbers of NK cells and CD56^dim^ and CD56^bright^ NK-cell subpopulations before and after HCMV reactivation (rct) or csCMVi events. Paired Wilcoxon test. (**D**) Representative flow cytometry plot showing the gating strategy for “memory-like” NK cells based on CD159c expression. (**E, G**) Kinetics of absolute numbers and frequencies of “memory-like” (CD159c^+^) NK cells^dim^ (CD56^dim^) (**F, H**) and mature (CD57^+^) “memory-like” NK cells^dim^ according to HCMV control, using two-group comparisons of controllers and csCMVi patients (**E-F**, Mann-Whitney U test) or three-group comparisons of controllers, no Rf-csCMVi, and Rf-csCMVi patients (**G-H**, Kruskal-Wallis). (**I-J**) Evolution of “memory-like” NK cells^dim^ (**I**) and mature “memory-like” NK cells^dim^ (**J**) counts between days 90 and 210 (top panels), or before and after HCMV reactivation/csCMVi events (bottom panels). Paired Wilcoxon test. *p < 0.05, **p < 0.01, ***p < 0.001. Abbreviations: alloSCT = allogeneic stem cell transplantation; csCMVi = clinically significant CMV infection; d = day; fcs = fold changes; rct = reactivation.

Next, we specifically focused on (NKG2C/CD159c^+^) “memory-like” NK-cell reconstitution (**Figure 5D**). Although not statistically significant due to considerable inter-individual variation, controllers consistently showed a trend toward higher “memory-like” CD56^dim^ (NK^dim^) NK-cell numbers (MMR 1.6-2.0) and frequencies (MMR: 1.2-1.4) than csCMVi patients (**Figure 5E**). Similarly, mature (CD57^+^) “memory-like” NK^dim^-cell counts tended to be higher in controllers than in csCMVi patients from day 210 onwards (**Figure 5F**).

We previously reported that low numbers and frequencies of “memory-like” NK cells were associated with severe and refractory csCMVi events^10^. To corroborate that trend in our current cohort, we again subdivided csCMVi patients into those with and without Rf-csCMVi. Indeed, patients with Rf-csCMVi had markedly lower numbers and frequencies of “memory-like” NK^dim^ cells and mature “memory-like” NK^dim^ cells compared to both no Rf-CMVi patients and controllers (**Figure 5G-H**).

Notably, controllers demonstrated significant expansion of “memory-like” NK^dim^ cells from day 90 to day 210 (16.0 vs. 35.8, median fold change=2.2, p=0.006) and from pre- to post-HCMV reactivation (9 vs. 50.0, median fold change=3.2, p=0.003). Although not reaching significance due to the smaller group size, similar changes were seen in non-Rf-csCMVi patients (day 90 to day 210: 8.9 vs. 36.3, median fold change=4.1, p=0.052; pre- to post-csCMVi: 24.2 vs. 36.3, median fold change=1.5, p=0.206). In contrast, median “memory-like” NK^dim^ -cell counts remained consistently <10 cells/μL in Rf-csCMVi patients (**Figure 5I**). Similar trends and differences between the groups were observed when specifically analyzing mature “memory-like” NK cells (**Figure 5J**).

In summary, consistent with prior evidence, controllers had higher numbers and frequencies of (mature) “memory-like” NK cells compared to csCMVi patients, especially those with Rf-csCMVi events. These differences increased over time and after reactivation events due to significant expansion of these cells in controllers and some patients with non-Rf-csCMVi, contrasting very limited expansion in those with Rf-csCMVi.

### HCMV-specific humoral IgG response correlates with the expansion of CD56^dim^ NK-cell subpopulations in controllers but not in csCMVi patients

Lastly, we sought to characterize associations between the reconstitution of the humoral immune response and HCMV-protective immune populations known to interact with IgG. Therefore, we analyzed correlations between HCMV-specific IgG levels and γδ T cells as well as “memory-like” NK cells.

Across all patients, correlations between HCMV-specific IgG levels and γδ T-cell subpopulations were generally weak and non-significant. However, the Vδ1^+^ γδ T-cell population showed a slight positive correlation with HCMV-specific IgG levels aggregated across all time points (rho=0.22, p=0.005, **Figure 6A–B**). Notably, this correlation was confined to the effector Vδ1^+^ γδ T-cell population, where weak to moderate correlations were observed both on aggregate across all time points (rho=0.38, p<0.001) and specifically on day 150 (rho=0.32, p=0.046) (**Figure 6A, C**), closely following the median day of HCMV reactivation (day 141) (**Figure 1B**). These trends were largely comparable among controllers and csCMVi patients.

**Figure 6.**
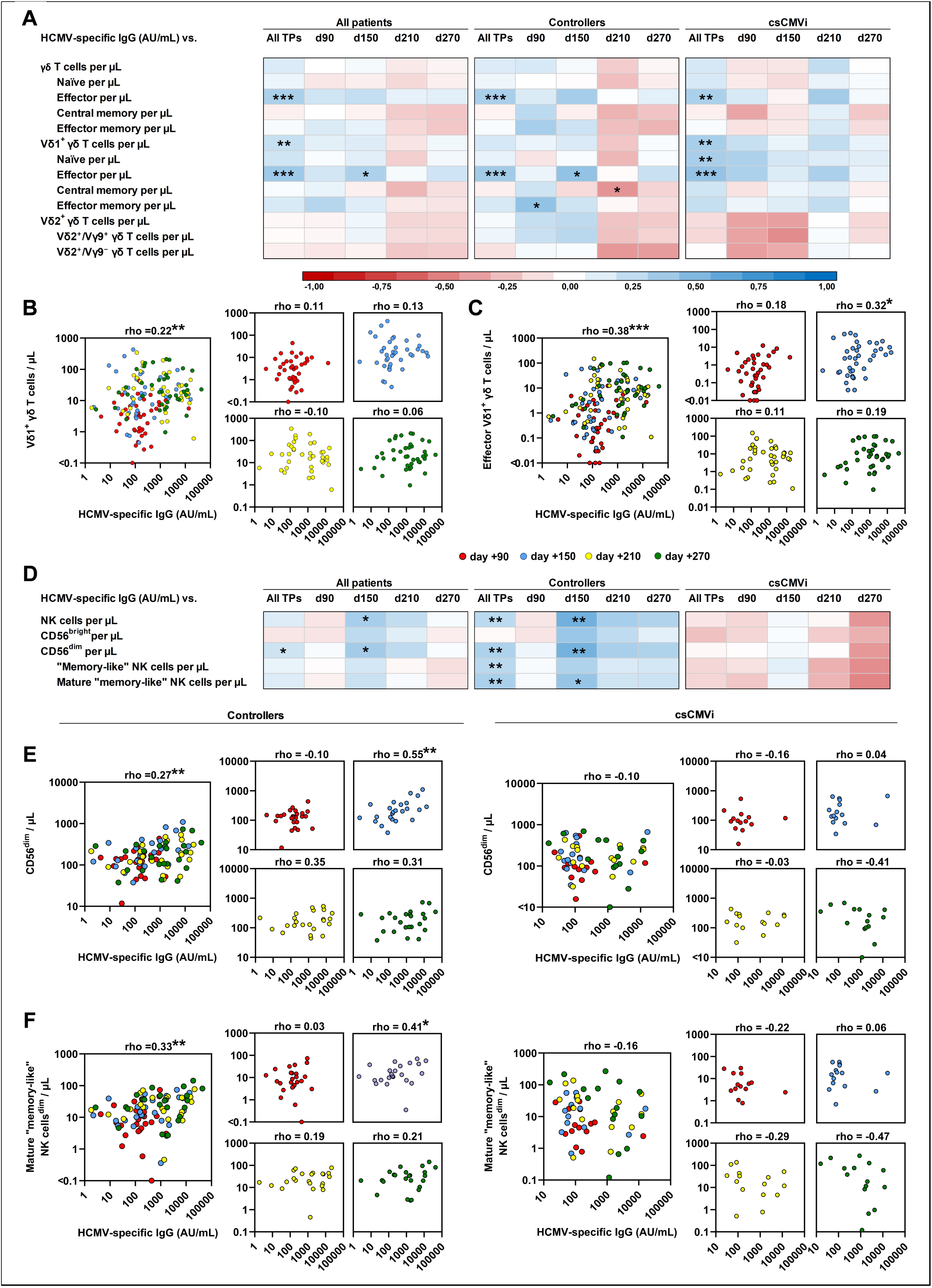
HCMV-specific humoral IgG response is associated with the expansion of CD56^dim^ NK-cell subpopulations in controllers but not in csCMVi patients. Correlations between HCMV-specific IgG and γδ T cells or NK cells were analyzed across all time points (TPs) and at individual time points. N = 40 patients. (**A**) Heatmap summarizing correlations of HCMV-specific IgG levels with numbers of γδ T cells and their subpopulations. (**B-C**) Correlation of HCMV-specific IgG with total Vδ1^+^ γδ T-cell (**B**) and effector Vδ1^+^ γδ T-cell (CD45RA^+^/CD27^−^) numbers (**C**) in all 40 patients across all time points (large panels) and at individual timepoints (small panels). (**D**) Heatmap summarizing correlations of HCMV-specific IgG levels with numbers of NK cells and “memory-like” NK cells. (**E-F**) Correlation of HCMV-specific IgG with total CD56^dim^ NK cells (**E**) and mature (CD57^+^) “memory-like” (CD159c^+^) CD56^dim^ NK cells (**F**) across all timepoints (large panels), and at individual timepoints (small panels). Data for HCMV controllers (left) and csCMVi patients (right) are displayed separately. All analyses were performed using Spearman’s rank correlation test. *p < 0.05, **p < 0.01, ***p < 0.001. Abbreviations: alloSCT = allogeneic stem cell transplantation; csCMVi = clinically significant CMV infection; HCMV = human cytomegalovirus; Ig = immunoglobulin; TPs = time points.

Interestingly, CD56^dim^ NK cells (rho=0.27, p=0.008), “memory-like” NK^dim^ cells (rho=0.27, p=0.007), and mature “memory-like” NK^dim^ cells (rho=0.33, p=0.001) showed significant correlations with HCMV-specific IgG levels on aggregate across all time points only in controllers but not in csCMVi patients. Positive correlation between HCMV-specific IgG levels and these cell populations in controllers was most pronounced on day 150, near the median time of HCMV reactivation (CD56^dim^: rho=0.55, p=0.005; “memory-like” NK^dim^ cells: rho=0.31, p=0.136; mature “memory-like” NK^dim^ cells: rho=0.41, p=0.046) (**Figure 6D-F**). In contrast, csCMVi patients showed an increasing disconnection between HCMV-specific IgG formation and memory-like” NK^dim^-cell reconstitution, resulting in a marked negative correlation by day 270 (mature “memory-like” NK^dim^ cells: rho=-0.47, p=0.076).

Overall, HCMV-specific IgG production correlated weakly with γδ T-cell reconstitution, with minimal differences between controllers and csCMVi patients. In contrast, CD56^dim^ NK-cell subpopulations, including (mature) “memory-like” NK cells correlated significantly with HCMV-specific IgG levels only in controllers. In contrast, a disconnection between these responses was seen in csCMVi patients.

## Discussion

Letermovir has revolutionized HCMV management in alloSCT recipients, yet late-onset HCMV reactivation after termination of letermovir prophylaxis is seen in 24.8 - 57% of patients^6–9^. Currently, there is no approved vaccine against HCMV^24^, and extension of letermovir prophylaxis from 100 to 200 days has not substantially reduced the incidence of csCMVi or improved overall survival^14^. This highlights the urgent need for alternative management strategies and better risk stratification tools. A key factor driving late-onset reactivation is the slow recovery of anti-HCMV immunity^11^. While T cells are well-recognized as central to HCMV immunity^10–12^, other key effectors, including HCMV-specific B cells, antibodies, “memory-like” NK cells, and γδ T cells^10^, have been scarcely studied.

Before the introduction of letermovir, early-onset HCMV reactivation commonly occurred before significant B-cell reconstitution could take place. In contrast, in the letermovir era, more readily reconstituted humoral immunity might play a more important role in controlling late-onset HCMV reactivation and disease. Indeed, HCMV-specific B cells were moderately reconstituted at the time of the first HCMV reactivation (median day 141) in our cohort, and significant further expansion of HCMV-specific humoral immunity was observed between days 150 and 210 in our cohort. This suggests that letermovir creates a window for B-cell reconstitution, in turn facilitating robust humoral responses following HCMV stimulation. Reactivation events in csCMVi patients drove the expansion of MBCs, with higher levels observed in csCMVi patients compared to controllers at the end of the observation period. This likely reflects a response to higher levels of viral replication followed by presentation of HCMV antigens. These prominent humoral responses underscore that future HCMV vaccines could leverage enhanced B-cell reconstitution, enabled by the shift to late-onset csCMVi in the letermovir era, to benefit alloSCT recipients.

Notably, HCMV-specific B cells displayed diverse antigenic specificities for three key antigens involved in viral dissemination: the fusion protein gB, the trimeric complex (gH/gL/gO), and the pentameric complex (gH/gL/UL128/UL130/UL131A)^24^. Following HCMV control in csCMVi patients, most MBCs were specific for gB and trimeric complex, whereas pentamer-specific MBCs were relatively scarce. These results align with prior clinical data suggesting that the gB/MF59 vaccine is one of the most effective HCMV vaccine candidate evaluated to date, reducing virus acquisition rates by 50% in seronegative individuals, with a strong correlation of gB-specific antibody titers and reduction in duration of viremia in solid organ transplant recipients^25,26^. The development of specific humoral immunity to multiple HCMV antigens after achieving viral control underscores the potential protective benefits of incorporating several well-defined antigens into vaccines and monoclonal antibody therapies. This approach is further supported by the observed reduction in HCMV viremia following treatment with a dual-antibody combination that targets both gH/gL and the pentameric complex^27,28^.

Furthermore, there was a noticeable difference in the timing of HCMV-specific IgG and IgM responses between controllers and csCMVi patients. Specifically, csCMVi patients exhibited a delayed onset of HCMV-specific IgG production and a stronger HCMV-specific IgM response than controllers. Consistent with this observation, Liu and colleagues reported that HCMV-specific IgG levels above 400 mg/dL at the time of letermovir cessation were associated with reduced risk of HCMV reactivation^9^. We hypothesize that the delayed switch may be driven by quantitatively and/or functionally impaired HCMV-specific Th-cell responses in patients developing csCMVi^10–12^. The importance of Th cells in class-switch recombination is well-documented, with deficiencies in Th and B-cell interplay linked to conditions such as Hyper-IgM syndrome. Future research exploring the interaction between HCMV-specific Th responses and humoral immunity could reveal strategies to enhance vaccine efficacy or develop targeted immunotherapies.

As a key component of long-term immunity, the humoral response provides sustained antibody production. However, effective viral control also relies on γδ T cells and memory-like NK cells. γδ T cells play a crucial protective role in preventing viral infections in alloSCT patients^29^. Both Vδ1+ and Vδ2+ γδ T-cell subpopulations possess protective capacities against infections^30^. In the context of HCMV, Vδ1+ γδ T cells play a protective role, whereas Vδ2+ γδ T cells appear to have limited involvement^30^, suggesting that a higher Vδ1+/Vδ2+ ratio may be more beneficial for HCMV immunity. In the pre-letermovir era, HCMV reactivation led to an expansion of γδ T cells, primarily driven by the Vδ2^−^ subpopulation, which consists predominantly of Vδ1^+^ cells^20,31^. The expansion of the Vδ2^−^ fraction resulted in a decreasing Vδ2^+^/Vδ2^−^ ratio^32^. Our data suggests that this trend persists even in alloSCT recipients receiving letermovir prophylaxis since both controllers and csCMVi patients showed an expansion of their Vδ2^−^ and Vδ1^+^ γδ T-cell populations following letermovir cessation. However, this expansion was more pronounced in csCMVi patients, resulting in an increased Vδ1^+^/Vδ2^+^ ratio after reactivation events compared to controllers. This may be attributed to the longer and more intense HCMV reactivation observed in csCMVi patients.

Interestingly, HCMV controllers already had elevated Vδ1^+^ γδ T-cell frequencies prior to letermovir cessation, which predicted successful HCMV control without progressing to csCMVi. This suggests a potential role for Vδ1^+^ γδ T cells in effective HCMV control. The protective role of Vδ1^+^ γδ T cells aligns with prior mechanistic studies demonstrating the capacity of Vδ1^+^ γδ T cells to lyse HCMV-infected cells and secrete antiviral cytokines such as IFNγ^20–22^. While the predictive performance of Vδ1^+^ γδ T-cell frequencies in our study would be insufficient for use as a standalone marker, quantifying these cells might be of interest for combinatorial immune-guided risk stratification. While analysis of (conventional) HCMV-specific T cells continues to predict future HCMV reactivation events in the letermovir era, our published data highlighted that incorporation of additional markers (e.g., reconstitution of regulatory T cells) might refine risk prediction models^11^. Additionally, the impact of letermovir prophylaxis on γδ T-cell reconstitution may have broader implications for alloSCT patients, given that γδ T cells are associated with reduced leukemia relapse and improved overall survival without increasing the risk of GvHD^29,33^.

HCMV infection not only alters γδ T-cell phenotypes but also profoundly reshapes the NK-cell repertoire, leading to the emergence of “memory-like” NK cells^34,35^. These cells have adaptive features, including clonal expansion, long-term persistence, and enhanced recall responses^23,36^. Their significant anti-HCMV effects are well-documented^23,36^ and are further supported by the reported association of low numbers of “memory-like” NK cells with an increased risk of future HCMV reactivation in alloSCT patients^37^. In our previous study, controllers showed higher frequencies and counts of “memory-like“

NK cells than csCMVi patients, particularly those with Rf-csCMVi^10^. This trend was corroborated in the present study. However, even patients with higher “memory-like” NK cell frequencies can develop csCMVi, suggesting that these cells alone are insufficient for viral control. This conclusion is supported by data from kidney transplant recipients, indicating that “memory-like” NK cells can suppress HCMV replication after its onset but cannot prevent its initiation^38^.

Both γδ T cells and “memory-like” NK cells are linked to the humoral immune response through their receptor CD16 (FcγRIII), an Fc receptor for IgG that mediates key immune functions such as ADCC^15–17^. Consequently, we investigated whether the more advanced humoral response in the letermovir era correlated with an expansion of these immune cell populations. We found that HCMV-specific IgG positively correlated with the expansion of effector γδ T cells in both controllers and csCMVi patients. CD16⁺ effector γδ T cells are known to expand following stimulation by stress-induced signals from HCMV-infected cells and persist long-term^21^, a process potentially supported by HCMV-specific IgGs. This likely enhances protection against HCMV, as HCMV-stimulated γδ T cells can produce IFNγ upon CD16 engagement with immunoglobulin-coated HCMV virions^15^.

The expansion of “memory-like” NK cells *in vitro* has been linked to polymorphic signal sequences of HCMV UL40 which can bind to HLA-E serving as a ligand for NKG-2C^39^. However, it appears that the most effective NKG-2C ligands of UL40 are relatively rare among clinical HCMV isolates (ref. Hammer et al), making our observation particularly intriguing that HCMV-specific IgG correlated positively with the expansion of CD56^dim^ NK-cell populations, including “memory-like” NK cells. A link between the pool size of “memory like” NK cells and HCMV-IgG was reported before in healthy seropositive individuals^40^ and was now seen in our study seen in alloSCT controllers but not in csCMVi patients. In contrast to CD56^bright^ NK cells, major parts of the CD56^dim^ subset strongly expresses CD16^41^. Notably, “memory-like” NK cells exhibit enhanced ADCC capacity and superior functionality compared to conventional NK cells, responding robustly to HCMV-infected cells in the presence of virus-specific antibodies^23^. Our finding of significant differences in the co-evolution of these cells and humoral immunity between controllers and csCMVi patients suggests that IgG-dependent “memory-like” NK cell-mediated anti-HCMV immunity may serve as a novel protective mechanism against csCMVi in the letermovir era, warranting further investigation in future studies.

The HCMV-specific IgG pool is highly heterogeneous^42^ and includes both virion neutralizing antibodies and other antibody subsets exhibiting diverse immune effector functions^43^. Identifying specific antibody subsets functionally associated with “memory-like” NK-cell and γδ T-cell responses might further clarify their protective roles. It could guide efforts to optimize both active and passive HCMV immunization strategies. For instance, recent studies have shown that presentation of the gB protein via mRNA induces stronger ADCC and broader gB-specific T-cell responses compared to adjuvanted protein vaccines^24^. Furthermore, while beyond the scope of this study, the use of functional assays might allow for more detailed characterization of the interplay between patient-derived specific IgGs and “memory-like” NK cells or γδ T cells in future studies.

This study has some limitations. Firstly, total HCMV-specific IgM and IgG levels were measured by a commercial chemiluminescent microparticle immunoassay without distinguishing neutralizing from non-neutralizing antibodies or those targeting specific HCMV antigens, which could provide deeper insights, as discussed above. Secondly, although we captured key time points in HCMV immune reconstitution, the 60-day intervals in our immune monitoring approach led to variable gaps between reactivation/csCMVi events and the corresponding pre- and post-event assessments. Thirdly, the use of cryopreserved PBMCs may have influenced phenotypic analyses. Future studies should address these limitations and incorporate multivariate analyses in larger cohorts to account for variables such as donor and recipient serostatus and immunosuppressive regimens.

Despite these limitations, our study highlights how the shift from early- to late-onset csCMVi driven by letermovir prophylaxis has led to an increased impact of humoral immune responses in anti-HCMV defense. We also provided further data supporting the role of “memory-like” NK cells as well as the first comprehensive analysis of γδ T-cell reconstitution in anti-HCMV immunity in the letermovir era. Vaccination strategies will become an increasingly important frontier in HCMV management in alloSCT recipients, with promising candidates like mRNA-1647 advancing to clinical trials (NCT05085366). Investigating the interplay between humoral and cellular immunity will be essential to guide the optimal approach and timing for vaccination and develop more effective therapeutic interventions such as antibody-based therapies. Moreover, early tracking of HCMV-specific B cells or Ig and/or analysis of γδ T-cell reconstitution might allow for fine-tuning prognostic immune monitoring strategies beyond assessing type-1 T-cell responses.

## Data Availability

The datasets generated and analysed in this study are available from the corresponding author upon reasonable request.

## Acknowledgments

This research was funded by a grant from the German Research Council (DFG) within the DFG Research Unit FOR 2830 (KR 5761/1-2, project number 398367752 (to S.K.); EI 269/10-2, project number 398367752 (to H.E.); DO 1275/7-2 (to L.D.); HE2526/9-2 (to H.H.)). S.K. was further supported by a fellowship of the Interdisciplinary Center for Clinical Research (IZKF) and the trust Forschung Hilft. The authors express gratitude to all participants for their contributions of blood samples. Special thanks are extended to Lubov Darst, Selina Grafelmann, and Anna Groß for their help in collecting patient samples, as well as Oana Butto for assistance with PBMC isolation. Minor parts of Figure 1A, and Figure 3A have been created with BioRender.com (https://BioRender.com/glnub6y). ChatGPT-4o (OpenAI) was used as a writing assistance tool for language editing and improving readability. The AI was used solely to enhance clarity and coherence, without contributing to the conceptualization, analysis, or interpretation of the study.

## Author Contributions

The study was conceived by C.D.L., H.E., S.W. and S.K. Patient enrollment and clinical documentation were performed by S.K. Experiments were planed and performed by C.D.L., H.G., K.K., B.W. and C.K. Data were analyzed by C.D.L., H.G., N.I., M.H., H.H, S.W. and SK. Data were visualized by C.D.L. Project administration and supervision were led by C.D.L, L.D., H.E., S.W. and S.K. Funding was acquired by L.D., H.E., and S.K. The original draft was written by C.D.L., S.W. and S.K. All co-authors reviewed, edited, and approved the manuscript.

## Conflict of Interest

The authors declare no conflicts of interest.

## Supplementary Figure Legends

**Figure S1.**
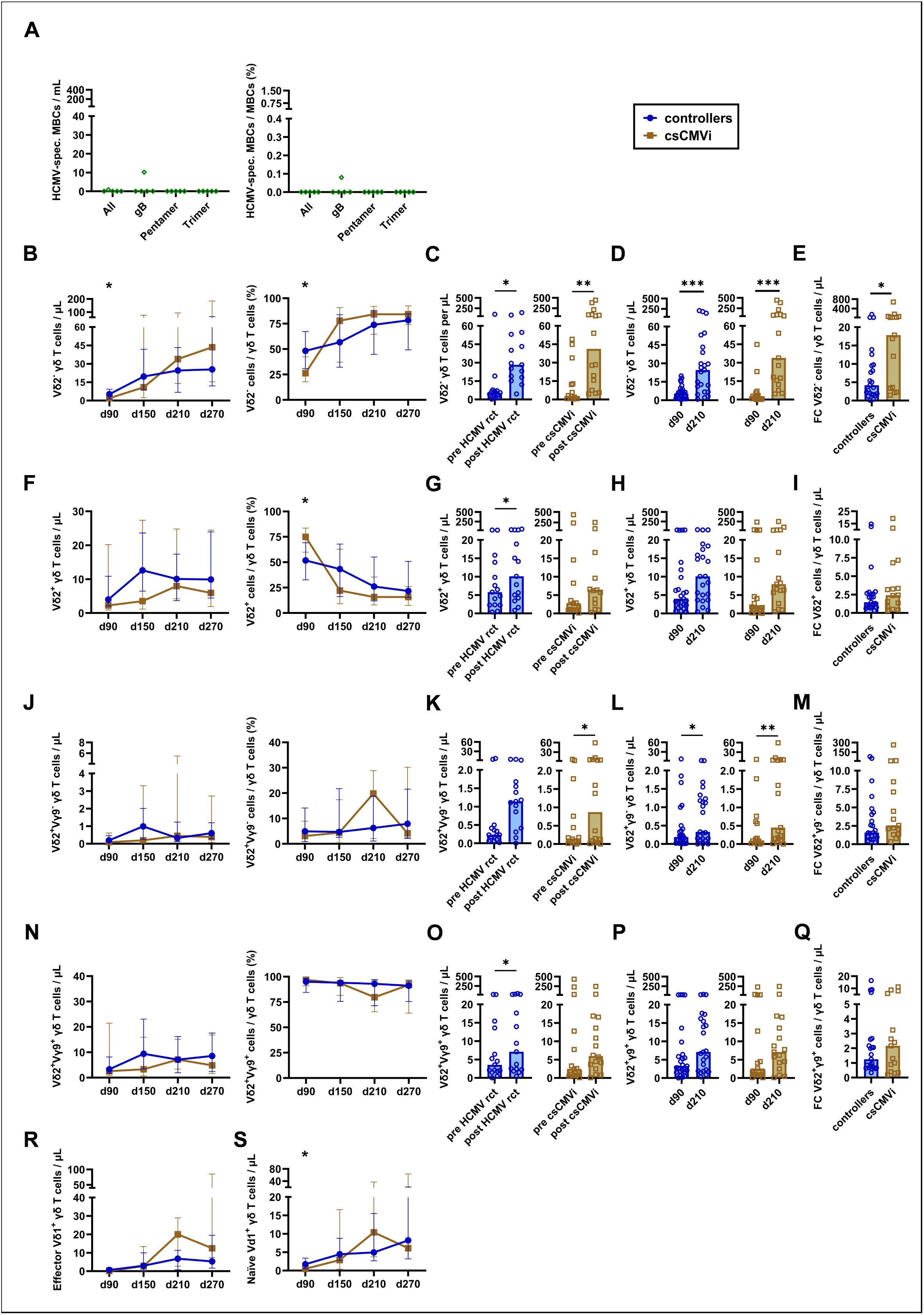
HCMV-specific B cells are undetectable in HCMV-seronegative patients, and HCMV reactivation is primarily associated with alterations in the Vδ2^−^ γδ T cell subset. **(A)** Absolute numbers and frequencies of HCMV-specific memory B cells (MBCs) targeting glycoprotein B (gB), the pentamer complex (gH/gL/UL128/UL130/UL131A), the trimer complex (gH/gL/gO), and all three antigens combined in HCMV-seronegative alloSCT recipients on day 270 (N = 5). **(B–Q)** Reconstitution of Vδ2⁻ (**B-E**), Vδ2⁺ (**F-I**), Vδ2⁺Vγ9⁺ (**J-M**), and Vδ2⁺Vγ9⁻ (**N-Q**) γδ T cells in 42 alloSCT recipients analyzed by flow cytometry. Absolute cell counts and frequencies are shown in **B**, **F**, **J**, and **N**. Mann-Whitney U test. Cell counts before and after HCMV reactivation or clinically significant CMV infection (csCMVi) are displayed in **C**, **G**, **K**, and **O**. Paired Wilcoxon test. Changes in cell numbers between day 90 and day 210 are shown in **D**, **H**, **L**, and **P**. Paired Wilcoxon test. Intra-individual fold changes (FC) from day 90 to day 210 are presented in **E**, **I**, **M**, and **Q**. Mann-Whitney U test). (**R-S**) Absolute cell counts of effector (**R**) and naïve (**S**) Vδ1⁺ γδ T cells were also analyzed (paired Wilcoxon test). *p < 0.05, **p < 0.01, ***p < 0.001.

## Supplementary methods

After thawing, global γδ T-, B-, and NK-cell phenotypes were analyzed, and HCMV-specific B cells were quantified using flow cytometry. For γδ T- and B-cell phenotyping, 5×10⁵ PBMCs per panel were stained using specific antibodies.

γδ T-cell staining consisted of TCR Vδ1 FITC, TCR γ/δ PE, CD27 PE-Vio615, TCR Vδ2 APC, TCR Vγ9 VioBlue, CD45RA VioGreen (Miltenyi), CD3 AF700 (BD), and Fixable Viability Dye eFluor780 (Invitrogen) in 90 µL Brilliant Stain Buffer (BD). B-cell staining utilized IgG FITC, CD27 PE-Vio615 (Miltenyi), IgM BV421, IgD BV605, CD20 BV650, CD21 Alexa Fluor700 (BioLegend), and Fixable Viability Dye eFluor780 in 90 µL Brilliant Stain Buffer.

For the NK-cell phenotype, 5×10⁵ PBMCs were washed in wash buffer (PBS + 1% FCS), blocked for 10 minutes at 4°C with 95 µL Brilliant Stain Buffer and 5 µL Blocking Reagent (Miltenyi), and stained for 30 minutes at 4°C in the same buffer. Antibodies used were CD159c PE-Vio770, CD57 APC (Miltenyi), CD56 BV510 BioLegend, CD3 AF700 (BD) and Fixable Viability Dye eFluor780.

HCMV-specific B cells were identified using biotinylated probes for the HCMV trimer (gH/gL/gO complex, The Native Antigen Company), pentamer (gH, gL, UL128, UL130, UL131A complex, The Native Antigen Company), and glycoprotein B (gB, SinoBiological). Proteins were biotinylated with EZ-Link Sulfo-NHS-Biotin (Thermo Scientific), incubated for 30 minutes at room temperature with a 20-fold molar excess of biotin reagent, and desalted using Zeba Spin Columns (Thermo Scientific). Biotinylated proteins were enriched with 15% glycerol (final concentration) and stored at 4°C. Before use, streptavidin PE and APC (Miltenyi) were bound to the probes, and 5×10⁶ PBMCs per protein/protein complex were stained. Additional antibodies used consisted of IgG FITC, CD27 PE-Vio615 (Miltenyi), IgM BV421, IgD BV605, CD20 BV650, CD21 AF700 (BioLegend), and Fixable Viability Dye eFluor780. Streptavidin BV510 (BioLegend) was used as a decoy probe to exclude non-specific B cells, with only double-positive (PE and APC) B cells considered specific.

Flow cytometric analysis was performed using a CytoFLEX cytometer and CytExpert v.2.4 software (Beckman Coulter). Data were analyzed in Kaluza v.2.1, and cell subpopulation frequencies were calculated by multiplying flow cytometry-derived percentages by absolute lymphocyte counts per µL of whole blood, as determined by clinical hematology.

### Gating strategy

#### B cells

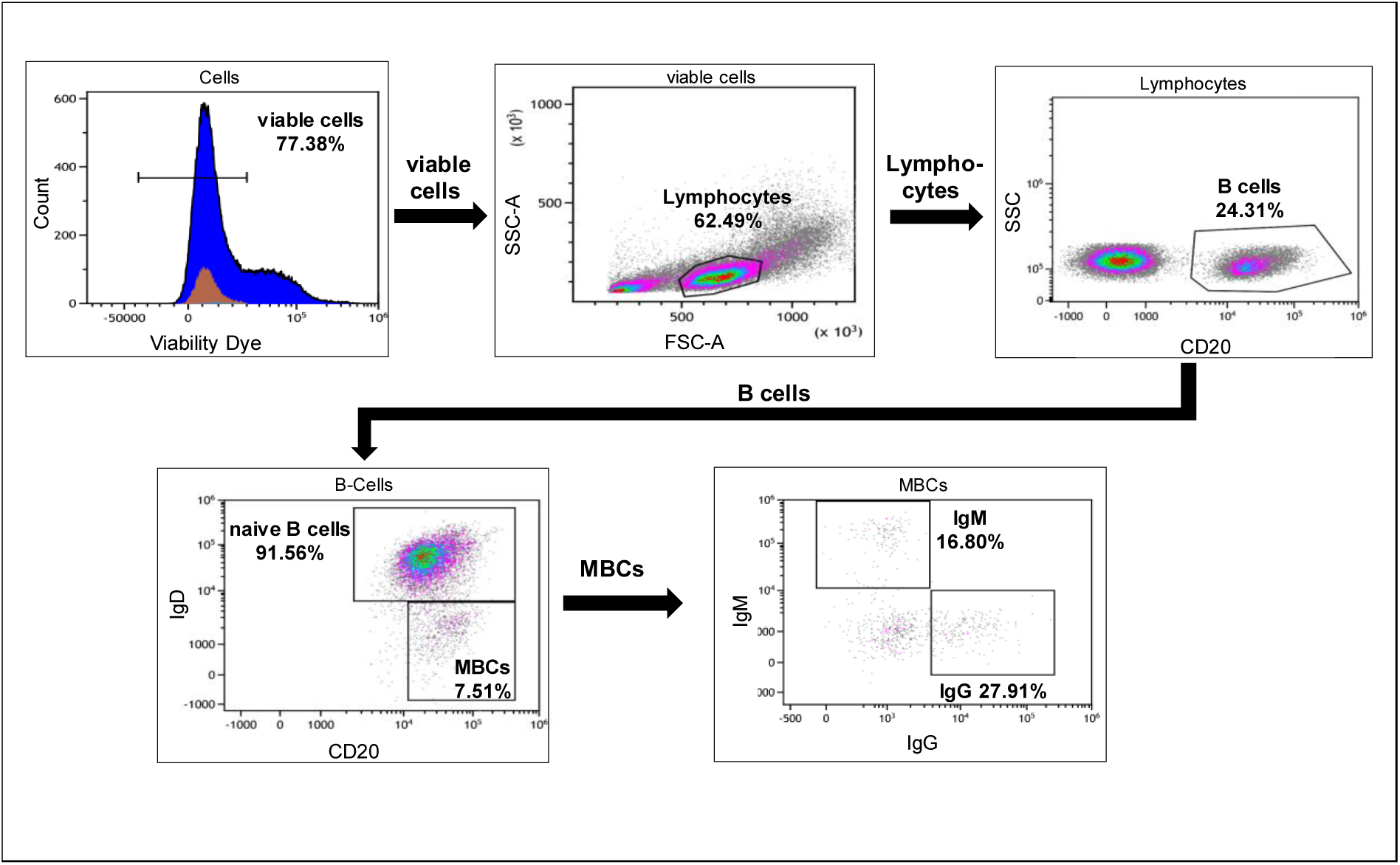

#### HCMV-specific memory B cells

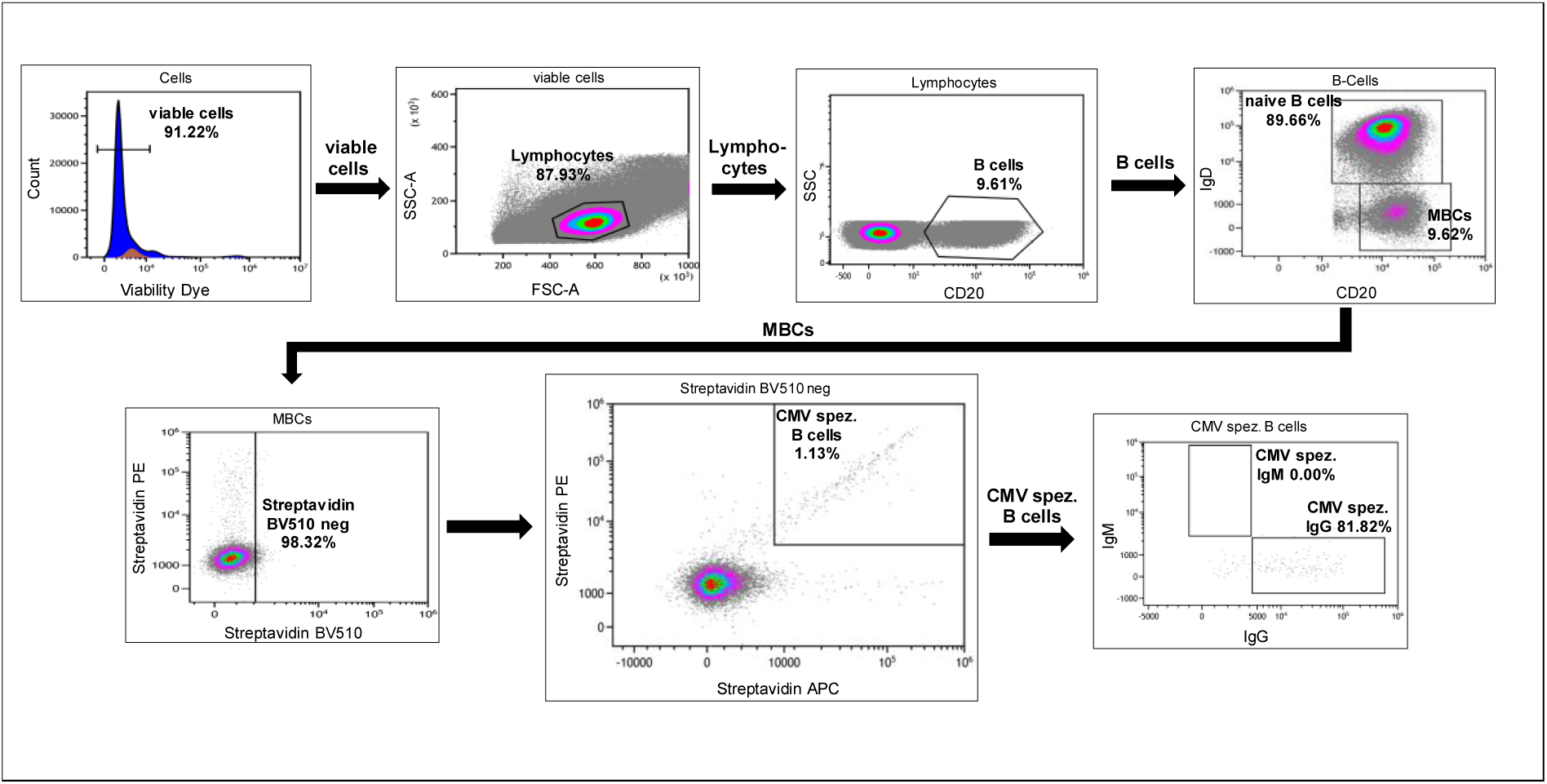

#### γδ T cells

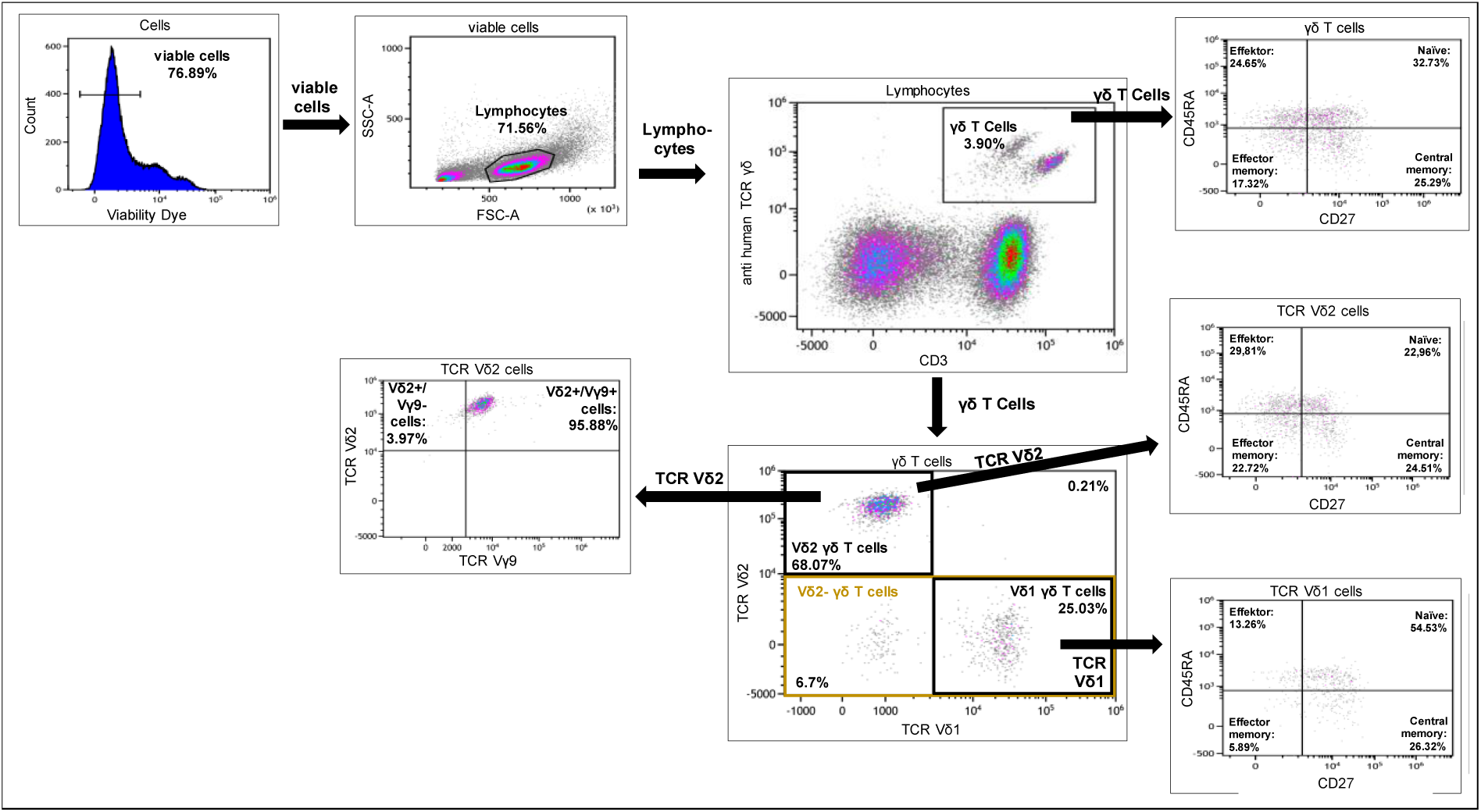

#### “Memory-like” NK cells

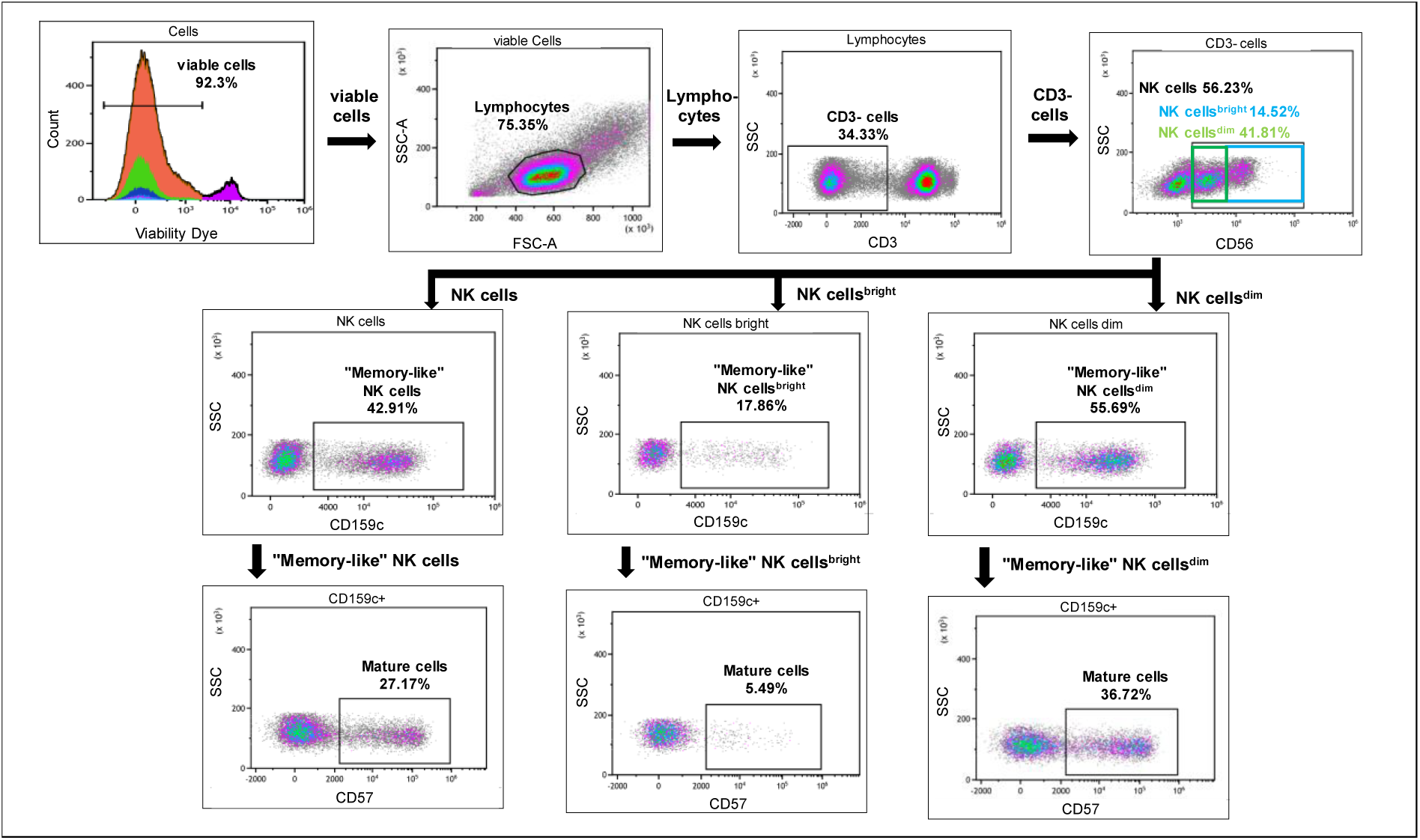

## Notes

### Competing Interest Statement

The authors have declared no competing interest.

### Author Declarations

This study was approved by the Ethics Committees of the University of Wuerzburg, Germany (protocol code 17/19-sc). Written informed consent was obtained from all patients.

